# Microbiome-based biomarkers to guide personalized microbiome-based therapies for Parkinson’s disease

**DOI:** 10.1101/2024.04.03.24305273

**Authors:** Haydeh Payami, Timothy R Sampson, Charles F Murchison, Zachary D Wallen

## Abstract

We address an unmet challenge in Parkinson’s disease: the lack of biomarkers to identify the right patients for the right therapy, which is a main reason clinical trials for disease modifying treatments have all failed. The gut microbiome is a new target for treatment of Parkinson’s disease, with potential to halt disease progression. Our aim was to develop microbiome-based biomarkers to guide patient selection for microbiome-based clinical trials. We used microbial taxa that have been robustly associated with Parkinson’s disease across studies and at high significance as dysbiotic features of Parkinson’s disease. Using individual-level taxonomic relative abundance data, we classified patients according to their dysbiotic features, effectively defining microbiome-based subtypes of PD. We show that not all persons with Parkinson’s disease have a dysbiotic microbiome, and not all dysbiotic Parkinson’s disease microbiomes have the same features. Grounded in robust and reproducible data from differential abundance studies, we propose an intuitive and easily modifiable method to identify the optimal candidates for microbiome-based clinical trials, and subsequently, for treatments that are personalized for each individual’s dysbiotic features. We demonstrate the method for Parkinson’s disease. The concept, and the method, is generalizable for any disease with a microbiome component.

## Introduction

Parkinson’s disease (PD) is the fastest growing neurologic disease in the world ^1^. PD is a progressively debilitating disease ^2^. The earliest manifestations are often constipation, sleep disorder, and hyposmia, leading to the cardinal movement disorders, and as disease progresses, most patients develop psychosis and dementia. Treatments are symptomatic. There have been many clinical trials for disease modifying treatment aimed at stopping disease progression, and all have failed (clinicaltrials.gov). PD is a highly heterogenous disease. One treatment will not work for all patients. Biomarkers to guide selection of the right patients for the right drug has been a high-priority, unmet need.

It has long been known that constipation, inflammation in the gut, and permeable gut membrane are common in PD and precede motor signs ^2^. Recent studies suggest some cases of PD start in the gut and spread to the brain, and in rodent models, PD pathologies are observed to spread from gut to brain ^3–7^. The gut origin raised the possibility that the gut microbiome could be involved in the brain-gut-axis pathology of PD. A healthy gut microbiome controls metabolism of drugs, toxicants and food, synthesis of vitamins and neurotransmitters, keeps the lining of the gut intact, protects against pathogens, maintains the proper functioning of the immune and nervous systems, and modulates brain-gut communication ^8^.

The gut microbiome is severely dysbiotic in PD, with characteristic features that are consistently observed in large-scale high-resolution shotgun metagenomic studies and meta-analysis of lower-resolution 16S studies ^9–13^. Differential abundance analyses, conducted in different geographic locations and with different methods, have converged on species and pathways that are commonly dysbiotic in PD, among them are the following which we chose for the present study: reduced levels of bacteria that degrade fiber and elevated levels of *Bifidobacterium*, *Lactobacillus*, opportunistic pathogens (e.g., *Porphyromonas*), *Escherichia coli*, and certain species of *Streptococcus* and *Actinomyces* ^9–13^. Functional analyses of these data suggest dysbiotic features of PD microbiome are relevant to multiple PD mechanisms, including increased abundance of pathogens and immunogens, decreased production of neuroactive molecules (dopamine, serotonin, glutamate and GABA), reduced capacity to degrade plant-based fiber which leads to short chain fatty acids deficiency, increased inflammation and compromised gut barrier, and elevated curli ^9^. Curli is a bacterial amyloid produced by *E.coli* which, in mice, induces alpha-synuclein aggregation, the hallmark of PD pathology ^14,15^. Data from genetic and toxicant-induced models of PD also suggest gut microbiome can trigger or contribute to various PD pathologies and the manifestation of motor and non-motor phenotypes ^16–19^, and that altering the microbiome via fecal microbiome transplantation (FMT)^20,21^, antibiotics treatments^22–25^, or high fiber diet^26^, each to some extent can reduce alpha-synuclein pathology, inflammation, and motor and non-motor dysfunction.

The gut microbiome is an emerging target for PD therapies ^27,28^. Results of early clinical trials with fiber supplementation ^29^ and FMT ^30–33^ are promising. Next-generation targeted therapeutics are forthcoming. One example (https://www.axialtx.com/our-programs/parkinsons-disease/) is a small molecule that reduces curli and hence could potentially attenuate disease progression by stopping curli-induced alpha-synuclein aggregation in the gut. The question is how to select patients to optimize success of microbiome-based clinical trials. Do all PD patients have elevated *E. coli* and curli? Is fiber supplementation sufficient to restore homeostasis to any dysbiotic microbiome? Are all persons with PD candidates for FMT? Here, we show that not all PD patients have a dysbiotic microbiome and not all dysbiotic microbiomes have the same features. We introduce microbiome-based biomarkers to guide patient selection for clinical trials, and upon success, as companion diagnostic for personalized treatment.

## Method

### Overview

We explored the possibility of subtyping PD patients based on the dysbiotic features of the microbiome; and using the microbiome profile of an individual to gauge their suitability for microbiome-based drug trials. The premise requires that disease associated features are already well established, i.e., reproduced across studies, using multiple differential abundance test methods controlling for confounders, at high statistical significance ^9–13^, and that the drug targeting a feature has undergone proper pre-clinical processes. The present paper describes a method to match patients to a treatment.

The overview of the method is shown in **Figure 1**. The concept and method are generalizable to any disorder with a microbiome component with well-established dysbiotic features, here, we focus on PD. The method requires a metagenomics reference dataset from a non-disease population. The reference dataset should be from the same population as the patients to be studied or treated (such reference datasets will soon be available for several populations). For demonstration, we select several features of PD as potential targets for treatment. (In reality, the feature can be a single gene or metabolite, a species, a cluster of related species, or multiple polymicrobial clusters that a drug has been developed to target.) We then calculate the mean relative abundance and confidence intervals (CI) of the features in the metagenomics dataset of non-disease population. We have now created the reference dataset. We define dysbiosis by setting thresholds for relative abundances in reference dataset (e.g., outside 95% CI). (We are cognoscente that “normal” and dysbiotic” microbiome are yet to be defined. Here, we use a working definition of dysbiotic to refer to abnormal levels of a well-established feature of PD microbiome). Patients can now be screened for dysbiotic features, which entails obtaining a stool sample, metagenomic sequencing and comparing the relative abundance of the features in the patient to the reference dataset and the thresholds that define dysbiosis.

**Figure 1.**
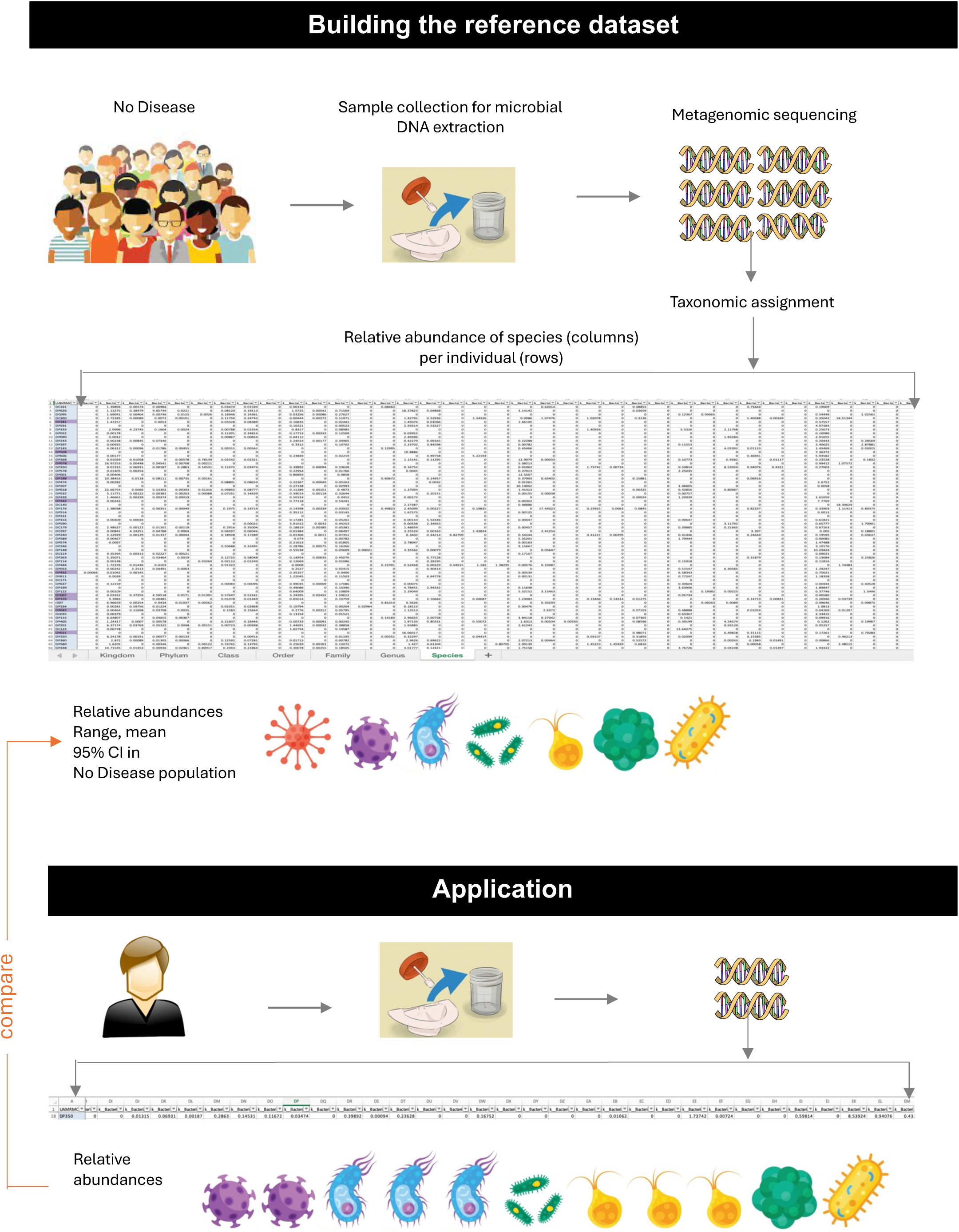
The method. For Parkinson’s disease, several disease-associated microbial features have already been well established, i.e., reproduced across studies at high statistical significance using multiple differential abundance test methods and including covariates and confounders ^9–13^. Clinical trials of microbiome-based treatments are underway. The present paper describes a method to match patients to treatments, excluding patients who do not have the feature of interest, and enriching the trial with patients with high abundance of the feature being targeted. A reference dataset is built by obtaining the metagenomic profiles of a non-disease cohort that represents the same population as the persons to be studied or treated. Once this infrastructure is in place, one can pick any feature (from a microbial gene to species to polymicrobial clusters), calculate its relative abundance and confidence interval (CI) in the non-disease group, and set thresholds for defining dysbiotic and non-dysbiotic. Application entails obtaining the metagenomic profile of the individual of interest and comparing it to the non-disease population. Here, we selected 6 PD-associated features (each a polymicrobial cluster), 5 of which are elevated in PD, and one reduced in PD ^9^. We set the threshold at 95%CI of the non-disease reference dataset. A person would be marked as dysbiotic for a feature if their relative abundance of that feature is above (for features that are elevated in PD) or below (for feature that are reduced) the 95%CI in non-disease population.

Species-level data and a step-by-step protocol to reproduce the current work are in the Supplement as well as on ASAP-ZENODO Workplace (https://zenodo.org/records/11126860). The source data we used is public, available for download as raw data from SRA or processed data from Zenodo. Metagenomic sequences (raw data) are available at NCBI SRA under BioProject ID PRJNA834801. The full taxonomic profiling data (processed data) is available at Zenodo (https://zenodo.org/records/7246185).

### Foundation of the method

Here, we use the data from the largest high-resolution metagenomic study of PD, published by Wallen et al. ^9^. The Wallen et al. study was compliant with all relevant ethical regulations and was approved by Institutional Review Boards at University of Alabama at Birmingham and by the Human Research Protection Office of United States Department of Defense (funding agency). All subjects signed informed consent, including granting permission to share and use their de-identified data in subsequent studies.

The dataset is composed of deep-shotgun next-generation metagenome sequences of microbial DNA, extracted from stool samples of 490 individuals with PD and 234 neurologically healthy controls (NHC). Each metagenome (data point) represents one sample taken from one individual. Wallen et al. conducted differential abundance test via a metagenome-wide association study (MWAS) and identified species, genes and pathways that are depleted or overly abundant in PD. The feature we use here have been reproduced in other metagenomics and meta-analyses ^9–13^. We also conducted correlations network analysis and identified polymicrobial clusters of species whose abundances grow or shrink together ^9^. The visual of PD microbiome in Wallen et al data is depicted in correlation network (**Figure 2**). Species are plotted according to correlation in their abundances, forming clusters of correlated species, using SparCC (RRID:SCR_022734, https://web.mit.edu/almlab/sparcc.html).

**Figure 2.**
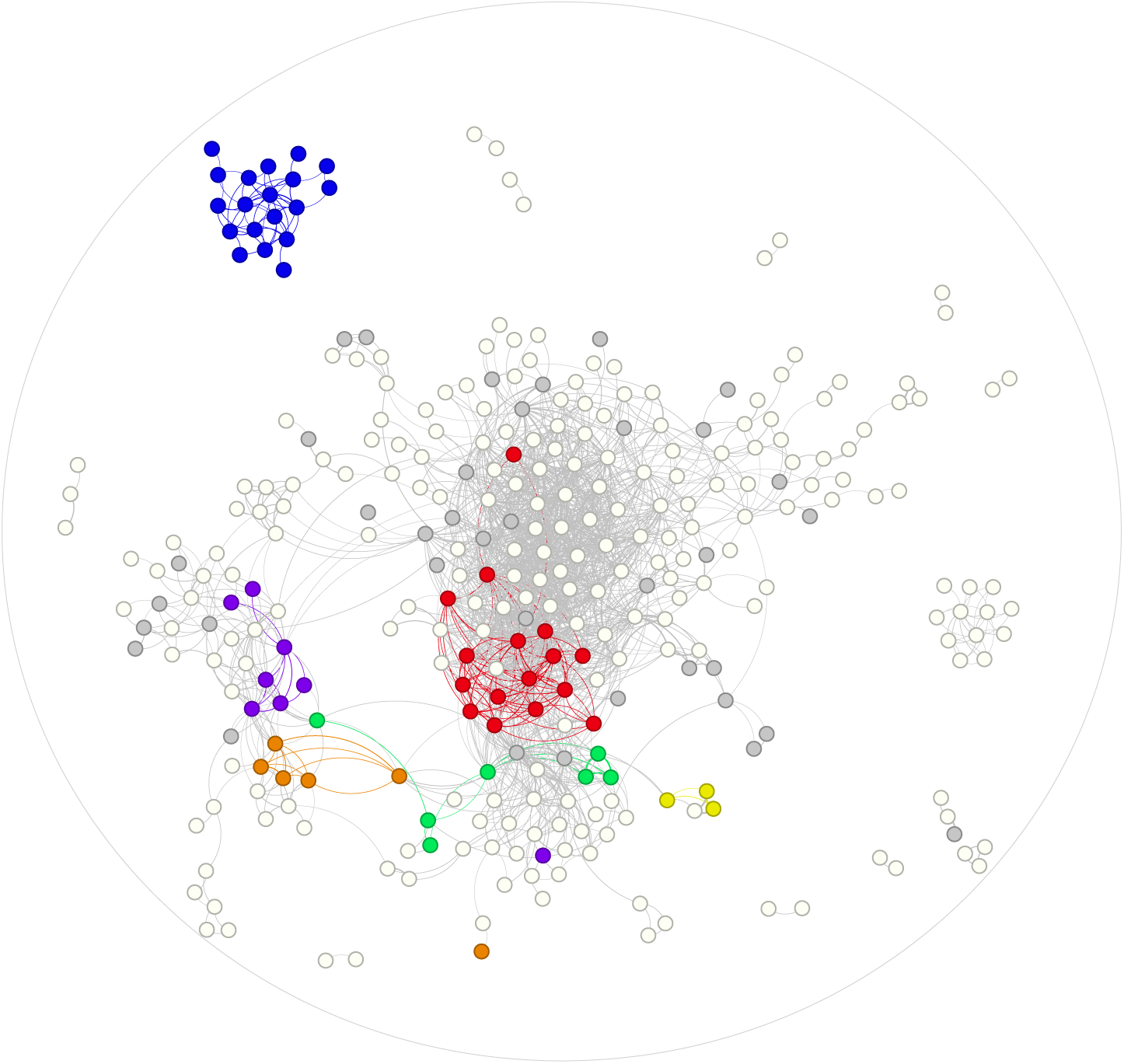
Polymicrobial clusters of species in PD gut microbiome. Pairwise correlation in relative abundances of all species detected in 490 unique PD gut metagenomes (i.e. 490 individuals with PD) was calculated and plotted (methods are described in Wallen et al ^9^). Each circle (node) represents one species and the curved lines (edges) connect species whose relative abundance correlate with each other (at correlation |r|>0.2, permuted P<0.05). Here, we have highlighted PD-associated species that were selected for this study, denoting by color the 6 dysbiotic features (species within each feature are listed in **Table 1**): Blue: Opportunistic pathogens (elevated in PD) Red: fiber-degraders (reduced in PD) Yellow: *E. coli* and *Klebsiella* (elevated in PD) Green: *Bifidobacteria* (elevated in PD) Orange: *Lactobacillus* (elevated in PD) Purple: *Streptococcus* and *Actinomyces* (elevated in PD) Grey: associated with PD, not selected as feature Empty circle: not associated with PD

### The concept

The published studies have been microbiome-centric, i.e., they identified dysbiotic features in aggregates of patients as compared to controls. We now turn the approach around and make it individual-centric, i.e., what does the microbiome of one patient look like? Is the microbiome dysbiotic in every PD patient? Are all dysbiotic features present together, or are some features present in a subset of patients, and if so, what percentage of patients has a given dysbiotic feature? The answers to these questions are fundamental to any microbiome-based therapeutic. To answer these questions, we had to (a) select features of interest that could potentially be a target of treatment and (b) create an operational definition of dysbiosis. The concept is generalizable to any disease and any microbiome in the body, and the method, both feature selection and definition of dysbiosis, can be modified to accommodate study design.

### Features of PD gut microbiome

For feature selection, we used the data reported in Wallen et al ^9^ and selected species that were significantly associated with PD (i.e., their relative abundance was elevated or reduced in PD not only in Wallen et al but reproduced across studies ^9–13^), plus the cluster of rare opportunistic pathogens were too rare to be tested individually but form a polymicrobial cluster that was shown in two independent datasets to be elevated in PD ^9,35^. We grouped the selected PD-associated species into six features based on the correlation in their abundances (|r|>0.2, P<0.05) and taxonomic relatedness (members of same genus). The six dysbiotic features (**Table 1**, **Figure 2**) are as follows: (1) the cluster of opportunistic pathogens (elevated in PD), (2) species of *Bifidobacteria* (elevated in PD), (3) species of *Lactobacillus* (elevated in PD), (4) *Streptococcus* and *Actinomyces* species that are elevated in PD, (5) *E. coli* and *Klebsiella species* that are elevated in PD, and (6) fiber-degrading bacteria (reduced in PD). There is more to the dysbiosis of PD gut than these six features. Here, the aim being demonstrating the method, we strived for a balance between minimizing complexity while capturing the most robust and biologically relevant features.

**Table 1.** Six main dysbiotic features of PD gut microbiome. We selected species that individually, or as a cluster, were significantly elevated or reduced in PD ^9–13^, and assigned them to a “feature” according to the correlation in their abundances (connectivity, see **Figure 2**) and their taxonomic relatedness (members of same genus). Here, our aim was to demonstrate the method, for which we strived for a balance between capturing the most robust association with PD and reducing the complexity. Features can be redefined to accommodate the aims of the investigation.

| Feature | Species composition of feature | PD association | Number of connections |
| --- | --- | --- | --- |
| Opp pathogens | *Actinomyces turicensis* | Increased as cluster | 8 |
| Opp pathogens | *Anaerococcus octavius* | Increased as cluster | 4 |
| Opp pathogens | *Anaerococcus vaginalis* | Increased as cluster | 5 |
| Opp pathogens | *Atopobium minutum* | Increased as cluster | 8 |
| Opp pathogens | *Corynebacterium amycolatum* | Increased as cluster | 2 |
| Opp pathogens | *Corynebacterium aurimucosum* | Increased as cluster | 2 |
| Opp pathogens | *Finnegoldia magna* | Increased as cluster | 7 |
| Opp pathogens | *Gemella asaccharolytica* | Increased as cluster | 2 |
| Opp pathogens | *Peptoniphilus harei* | Increased as cluster | 1 |
| Opp pathogens | *Peptoniphilus lacrimalis* | Increased as cluster | 3 |
| Opp pathogens | *Peptoniphilus* sp HMSC062D09 | Increased as cluster | 7 |
| Opp pathogens | *Porphyromonas asaccharolytica* | Increased | 6 |
| Opp pathogens | *Porphyromonas* sp HMSC065F10 | Increased as cluster | 11 |
| Opp pathogens | *Porphyromonas uenonis* | Increased as cluster | 6 |
| Opp pathogens | *Prevotella bergensis* | Increased as cluster | 1 |
| Opp pathogens | *Prevotella buccalis* | Increased as cluster | 10 |
| Opp pathogens | *Prevotella disiens* | Increased as cluster | 1 |
| Opp pathogens | *Prevotella timonensis* | Increased as cluster | 8 |
| Opp pathogens | *Varibaculum cambriense* | Increased as cluster | 10 |
| *Bifidobacteria* | *Bifidobacterium dentium* | Increased | 12 |
| *Bifidobacteria* | *Bifidobacterium bifidum* | Increased | 3 |
| *Bifidobacteria* | *Bifidobacterium breve* | Increased | 24 |
| *Bifidobacteria* | *Bifidobacterium gallinarum* | Increased | 14 |
| *Bifidobacteria* | *Bifidobacterium longum* | Increased | 5 |
| *Bifidobacteria* | *Bifidobacterium pullorum* | Increased | 12 |
| *Bifidobacteria* | *Bifidobacterium saeculare* | Increased | 13 |
| *Lactobacillus* | *Lactobacillus fermentum* | Increased | 14 |
| *Lactobacillus* | *Lactobacillus gasseri* | Increased | 10 |
| *Lactobacillus* | *Lactobacillus paragasseri* | Increased | 9 |
| *Lactobacillus* | *Lactobacillus salivarius* | Increased | 9 |
| *Lactobacillus* | *Lactobacillus reuteri* | Increased | 1 |
| *Lactobacillus* | *Lactobacillus rhamnosus* | Increased | 11 |
| *Strepto / Actino* | *Actinomyces naeslundii* | Increased | 4 |
| *Strepto / Actino* | *Actinomyces oris* | Increased | 16 |
| *Strepto / Actino* | *Actinomyces* sp HPA0247 | Increased | 3 |
| *Strepto / Actino* | *Actinomyces* sp oral taxon 448 | Increased | 3 |
| *Strepto / Actino* | *Streptococcus anginosus* group | Increased | 10 |
| *Strepto / Actino* | *Streptococcus mutans* | Increased | 13 |
| *Strepto / Actino* | *Streptococcus vestibularis* | Increased | 15 |
| *Strepto / Actino* | *Streptococcus lutetiensis* | Increased | 3 |
| *E. coli / klebsiella* | *Escherichia coli* | Increased | 5 |
| *E. coli / klebsiella* | *Klebsiella pneumoniae* | Increased | 3 |
| *E. coli / klebsiella* | *Klebsiella quasipneumoniae* | Increased | 3 |
| Fiber-degraders | *Blautia wexlerae* | Decreased | 17 |
| Fiber-degraders | *Blautia hansenii* | Decreased | 17 |
| Fiber-degraders | *Anaerostipes hadrus* | Decreased | 48 |
| Fiber-degraders | *Clostridium* sp CAG 58 | Decreased | 27 |
| Fiber-degraders | *Eubacterium eligens* | Decreased | 47 |
| Fiber-degraders | *Eubacterium hallii* | Decreased | 52 |
| Fiber-degraders | *Eubacterium ramulus* | Decreased | 25 |
| Fiber-degraders | *Eubacterium rectale* | Decreased | 21 |
| Fiber-degraders | *Eubacterium* sp CAG 38 | Decreased | 22 |
| Fiber-degraders | *Faecalibacterium prausnitzii* | Decreased | 59 |
| Fiber-degraders | *Fusicatenibacter saccharivorans* | Decreased | 45 |
| Fiber-degraders | *Roseburia faecis* | Decreased | 21 |
| Fiber-degraders | *Roseburia intestinalis* | Decreased | 21 |
| Fiber-degraders | *Roseburia inulinivorans* | Decreased | 27 |
| Fiber-degraders | *Ruminococcus bicirculans* | Decreased | 33 |
| Fiber-degraders | *Ruminococcus lactaris* | Decreased | 12 |

### Definition of dysbiotic and non-dysbiotic

Defining healthy and dysbiotic microbiome has been a challenge. For this project, we limited the definition of dysbiotic to a metagenome that has very high or very low relative abundance of a feature that is already well-established to be elevated or reduced in PD. To define very high and very low, we set extreme thresholds to maximize the odds that the patient being enrolled in trial is an optimal candidate. These thresholds can be changed to accommodate study design, or relaxed when a drug is approved, and the biomarker becomes a companion diagnostic for treatment.

For each feature, and per person, we calculated total relative abundance of each feature by summing the relative abundances of the species within it. We then calculated the mean relative abundance and its 95% CI in NHC (reference dataset). Next, we turned to individual-level PD data. Considering each feature separately, we called a PD person’s metagenome dysbiotic if the relative abundance of the feature was outside the 95% CI of NHC. For features that are elevated in PD in aggregate data, a PD individual’s gut was called dysbiotic if the relative abundance of the feature in that person was greater than the upper bound of 95% CI of NHC. For the feature that is reduced in PD, a PD individual’s microbiome was called dysbiotic if the relative abundance of the feature in that person was less than the lower bound of 95% CI of NHC (**Table 2**).

**Table 2.** A working definition of dysbiotic for identifying microbiome-based subtypes of PD. In this context, the term dysbiotic refers to a microbiome in which the relative abundance of a well-established PD-associated feature is very high (for features that are elevated in PD) or very low (if reduced in PD). Here, each feature is composed of several PD-associated species, as shown in Table 1. The relative abundance of each species in each individual was calculated as reported before ^9^ and can be found here https://zenodo.org/record/7246185. We calculated relative abundance of each feature in each neurologically healthy control (NHC) and individual with PD by summing the relative abundances of the species within the feature for that individual. We then calculated the mean and 95% confidence interval (CI) of the mean in the NHC and PD separately. We set the threshold for defining dysbiotic at the upper 95% CI of NHC for features that are elevated in PD, and at lower 95% CI of NHC for fiber-degraders that are reduced in PD. Last column is the number of PD patients, among 490 total, who were called dysbiotic for each feature. For example, in 161 of 490 PD patients, the relative abundance of *Bifidobacteria* was higher than 2.66% which was the upper 95% CI limit of NHC. Similarly, in 333 of 490 patients, the relative abundance of fiber-degrading bacteria was lower than 20.81% which was the lower 95% CI limit in NHC. See Figure 3 for overlaps.

| Feature | Relative abundance of feature in 234 NHC |  |  | Relative abundance of feature in 490 PD |  |  | N among 490 PD outside 95% CI of NHC |
| --- | --- | --- | --- | --- | --- | --- | --- |
|  | Mean | Upper 95% CI | Lower 95% CI | Mean | Upper 95% CI | Lower 95% CI |  |
| Opp. pathogens | 0.01% | **0.01%** | 0.00% | 0.10% | 0.20% | 0.00% | 66 (13%) |
| *Bifidobacteria* | 2.06% | **2.66%** | 1.47% | 5.07% | 5.98% | 4.17% | 161 (33%) |
| *Lactobacillus* | 0.11% | **0.19%** | 0.04% | 0.55% | 0.74% | 0.36% | 103 (21%) |
| *Streptococcus / Actinomyces* | 0.19% | **0.33%** | 0.06% | 0.39% | 0.54% | 0.25% | 94 (19%) |
| *E. coli / Klebsiella* | 1.47% | **2.23%** | 0.72% | 1.94% | 2.44% | 1.43% | 86 (18%) |
| Fiber-degraders | 22.35% | 23.88% | **20.81%** | 15.29% | 16.36% | 14.22% | 333 (68%) |

A key question was whether all individuals with PD have a dysbiotic gut microbiome. To answer this question, we needed an operational definition of non-dysbiotic. We defined non-dysbiotic per feature as being well within the norm of NHC range, i.e., below the mean of NHC if the feature is elevated in PD, and above the mean if a feature is reduced in PD. When considering all six features, a PD person was designated as not having a dysbiotic microbiome only if their relative abundance for the five elevated features were all at or below the mean of NHC, and their relative abundance for reduced fiber degrading feature was at or greater than the mean in NHC.

## Results

### Not all PD metagenomes are dysbiotic, and not all dysbiotic microbiomes have the same features

In Wallen et al data, among the 490 individuals with PD studied, 392 (80%) had at least one dysbiotic feature (**Figure 3a**). 98 (20%) did not have any dysbiotic feature defined as being outside the 95% CI of NHC. Using the more cautious definition of non-dysbiotic, i.e. elevated features being at or below the NHC mean and reduced features at or above the NHC mean, 76 (16%) of PD microbiomes were not dysbiotic. The patients without a dysbiotic microbiome can be identified and excluded from microbiome-based therapeutics; doing so will boost power and odds of success of clinical trials, and in clinical setting, will spare these patients unnecessary trial and error in finding the right treatment.

**Figure 3.**
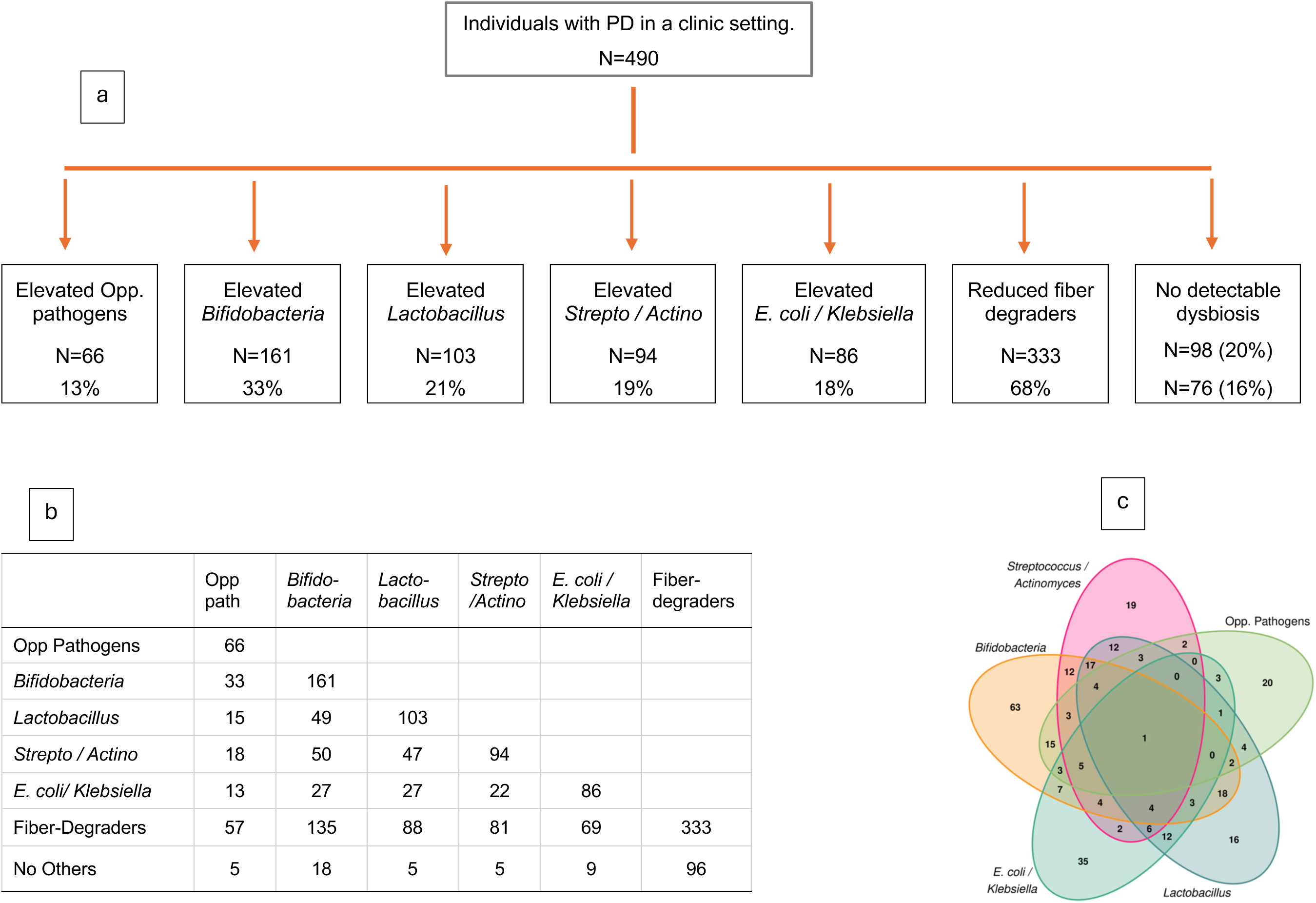
Subtyping PD by the dysbiotic features of the gut microbiome. The total number of patients is 490. The total number of patients with a dysbiotic feature is 392 (=490-98). The number of patients with a given dysbiotic feature are given in a-c. Actual numbers are given so that percentages can be calculated with the desired denominator.

a. Proportion of PD patients with different dysbiotic features.
  ▪ Elevated features: number of PD persons, among the 490, who had an abundance higher than upper limit of the 95% CI of the abundance in neurologically healthy control (NHC).
  ▪ Reduced feature: number of PD persons, among the 490, who had an abundance lower than the lower limit of 95% CI of the abundance in NHC.
  ▪ No detectable dysbiosis: 98 of 490 (20%) did not fall outside 95% CI of NHC for any feature. 76 of 490 (16%) of PD persons fit the more cautious definition of non-dysbiotic, whose abundances for every elevated feature (opportunistic pathogens, *Bifidobacteria*, *Lactobacillus*, *Streptococcus* and *Actinomyces* (Strepto/Actino), and *E. coli/Klebsiella*) was below the NHC mean, and their abundance for reduced fiber-degrading bacteria was above the NHC mean.
b. Pairwise overlap in features, n=number of patients, from 490, who have the two specified features. The total for six features (the diagonal) is 843, nearly double the number of 490 subjects, because features do co-occur in an individual. For example, 66 of 490 subjects had elevated (>95%CI of NHC) levels of opportunistic pathogens, among them, 33 (50%) also had elevated Bifidobacteria. Or the example in the text, 71% (=(333-96)/333) of low fiber-degraders had high level of at least one other feature.
c. Overlap across the 5 features that are elevated in PD (fiber-reducers not included for legibility). For example, 19 subjects have elevated levels (>95%CI of NHC) of *Streptococcus* and *Actinomyces* only, another 2 subjects have high *Streptococcus* and *Actinomyces* and high opportunistic pathogens, another 3 have high *Streptococcus* and *Actinomyces,* high opportunistic pathogens, and high *Lactobacillus*, etc.

Severe reduction in fiber-degrading bacteria was seen in 333 of 490 of PD patients (68%) (**Figure 3a**). Among the 333 low fiber-degraders, only 96 had no other dysbiotic features (**Figure 3b**), hence 71% (=(333-96)/333) of patients with low fiber-degraders had high levels of at least one other dysbiotic feature. Thus, while deficiency of fiber degraders is very common, only 20% (=96/490) of PD patients had low fiber-degraders as their sole feature. This suggests that while fiber treatment would be beneficial for majority of patients, a subset would stand to benefit the most from fiber, while others may need treatment for their other dysbiotic features as well.

Features that are elevated in PD were found in subsets of patients: high levels of opportunistic pathogens was detected in 66 (13%) of the 490 PD, *Bifidobacterium* in 161 (33%), *Lactobacillus* in 103 (21%), *Streptococcus* and *Actinomyces* in 94 (19%), and *E. coli* and *Klebsiella* in 86 (18%). To demonstrate utility of the biomarker, consider *E. coli* which has been linked to pathobiology of PD. *E. coli* encodes curli, an amyloid that induces alpha-synuclein aggregation in mice ^14,15^. Alpha-synuclein aggregation is a pathogenic hallmark of PD. Moreover, *E. coli* and genes that encode and regulate curli production are elevated in PD gut ^9^. A drug developed against *E. coli* or curli will likely have a better odds of success in clinical trial if patients were chosen from the 18% that have high levels of *E. coli* and curli.

There is some overlap across the groups shown in **Figure 3a**, because these features are not mutually exclusive. For example, as shown in **Figure 3b**, among the 66 patients with high opportunistic pathogens, 33 (50%) also had high *Bifidobacteria*, 15 (23%) had high *Lactobacillus*, 18 (27%) had high *Streptococcus* and *Actinomyces*, 13 (20%) had *E. coli* and *Klebsiella*, and 57 (86%) had low fiber degraders. These data suggest that at the present time, with fiber and FMT being the only microbiome-based treatments, the most rational option for treating the patients who have a severely dysbiotic microbiome is cleansing the microbiome with antibiotics followed by FMT, excluding those without dysbiosis and those with only low fiber-degraders. We are not advocating for FMT or any treatment modality per se, rather, pointing out the utility of biomarkers in designing a study as well as choosing the patients to study and treat.

### Application of the biomarker tool

Defining the features of interest and setting abundance thresholds to define dysbiotic/non-dysbiotic in a healthy population will constitute the reference dataset for this biomarker tool. To determine the composition of the metagenome of a patient will entail obtaining a stool sample, extracting the DNA, sequencing, bioinformatic processing and taxonomic assignment of sequences, and calculating relative abundances of the microbial features in the individual’s metagenome. The patient’s relative abundances for the features of interest are then compared to the reference dataset to determine if they are dysbiotic (**Figure 1**).

## Discussion

Meta-analyses and large metagenomic studies have converged on certain microbial features that consistently and significantly are altered in PD microbiome, independent of geography, confounding covariates, and methodology. These well-established disease-associated taxa are pre-requisite to, and the foundation of biomarkers described here. Here, we used these robust associations to define subtypes of PD based on dysbiotic features of microbiome. We introduced a microbiome-based biomarker tool to guide patient selection for clinical trials and to serve as companion diagnostic for personalized treatments. With this tool, we can “see” if and what is dysbiotic in an individual and act accordingly. The method is applicable for any patient at any stage of disease. The tool can be easily modified (by redefining features and adjusting thresholds) as the field advances, and to meet varying needs of different studies. Here, we demonstrated the development and the potential utility of the tool for identifying subtypes of PD based on their microbiome features. The concept is generalizable to any disease with a microbiome component.

Treating a dysbiotic microbiome can potentially be disease modifying, which has been an elusive goal in PD field. Functional inference of human data and experiments in mice indicate dysbiotic features of PD microbiome contribute to several disease mechanisms, including alpha-synuclein aggregation which is at the core of PD pathology, disrupted neuro-signaling, and inflammation ^9,15,16^. It is therefore reasonable to work towards microbiome-based therapies that could halt disease progression. As we learn which features are causal and which result from disease (an active area of research), microbiome-based biomarkers will be useful for identifying pathogenic dysbiosis early to prevent disease before onset.

Microbiome-based treatments under consideration for PD include prebiotics (e.g., fiber supplements), probiotics (beneficial bacteria packaged and marketed direct to consumer), FMT, and next generation therapeutics that are targeted to a specific feature (e.g., small molecules, phage, CRSPR-Cas9). The microbiome-based biomarkers described here can serve as companion diagnostic for all treatment modalities. Take fiber supplementation for example, which is safe and likely beneficial for everyone. We show here that indeed, the majority of PD patients have severely depleted levels of fiber-degrading bacteria. Increasing fiber intake is effectively feeding these bacteria and causing them to grow and multiply, and it has shown promise in an early clinical trial with newly diagnosed PD patients ^29^. The value of coupling biomarkers with fiber treatment is in identifying patients who, in addition to being low on fiber-degraders, have elevated levels of harmful pathogenic bacteria, which is an important consideration both for patient selections for clinical trials and for treating the individuals. As for probiotics on the market, most are composed of “beneficial” species of *Bifidobacteria* and/or *Lactobacillus*, which ironically, are highly elevated in PD. The value of the biomarker in this case is to avoid supplementation that is counter indicated. FMT is showing promise for PD ^30^, but it carries risks concerning safety and compatibility of donor sample with host environment. Using biomarkers, unnecessary FMT can be avoided, sparing the patients who do not have a dysbiotic microbiome and those whose only problem is low fiber-degraders. Targeted treatments could replace the need for FMT, and they too would benefit from a companion diagnostic. For example, for a treatment targeted to abolish the opportunistic pathogens, the biomarker can identify the 13% of patients who would be the best candidates.

Currently, we do not know if alleviating one dysbiotic feature would restore homeostasis to the rest of the microbiome, and what that central feature might be. Consumption of fiber is the safest and simplest treatment. In a clinical trial performed on 20 newly diagnosed PD patients, treatment with fiber increased the abundance of fiber-degrading bacteria and production of short chain fatty acids and resulted in shifts in the microbiome community ^29^. Fiber-degrading species are in the center of the correlation network with tight positive and negative correlations to several PD-associated species (**Figure 2**). It is therefore plausible that fiber treatment, aimed to increase the abundance of fiber-degrading bacteria, may restore equilibrium to PD-associated species that map to the center of network and are either positively or negatively correlated with fiber-degraders. However, while possible, it is less likely that fiber treatment alone would eliminate overabundance of opportunistic pathogens, for example, considering that opportunistic pathogens are a dense isolated cluster with no connection to fiber-degrading species.

Profiling the patients’ metagenome and comparing them to the reference dataset will show where they fall in the spectrum of dysbiosis. This concept, and the proposed method, holds true regardless of feature, disease, or dataset. However, the relative abundances of the features and therefore the range and thresholds for defining dysbiosis may differ across populations. Hence, it is important that the reference dataset is from the same source population as patients. Here, we used a cohort of neurologically healthy older adults from Birmingham, Alabama in United States for our reference dataset, as our patients are also from Birmingham. There are regional variation in microbiome, reflecting cultural, dietary, socioeconomic, and geographic influences. Despite regional variations, the main PD-associated features are robust and reproducible ^9,11,12,35^. Even so, the relative abundances, and hence the thresholds for defining dysbiosis, may still vary across populations. Not every investigator can amass hundreds of controls to investigate population-specific abundances to set their thresholds. This impediment will be overcome soon by current efforts to collect all data generated and made public. These collaborative global efforts are aimed at meta-analyses, a by-product of which will be open-access to well-curated datasets from across the world that can be used as reference datasets to generate population-specific thresholds for biomarkers and companion diagnostics.

## Data availability

The dataset used for biomarker development is publicly available, open access with no restrictions, as described in detail in Wallen et al 2022 ^9^. The dataset is available on two repositories to enable the investigators to download the data in either the raw or processed form. The raw metagenomic sequences and accompanying metadata are on NCBI SRA under BioProject ID PRJNA834801 [https://www.ncbi.nlm.nih.gov/bioproject/834801]. The post-QC and post taxonomic profiling data are on Zenodo [https://zenodo.org/record/7246185]. Here we used species-level data (presented in Supplement) that was extracted from the taxonomic profiling data downloaded from Zenodo in January 2024.

## Code and software

Algorithm was developed using Microsoft Excel v.16.84 (RRID:SCR_016137) https://www.microsoft.com/en-gb/. Numbers were double checked using R v4.3.3. (RRID:SCR_001905) https://www.r-project.org/. Both Excel and R code are provided in the Supplement, as well as in ASAP-ZENODO Workplace (https://zenodo.org/records/11126860).

## Acknowledgements

This work was supported by The Strain Endowed Chair in Parkinson’s Disease to the University of Alabama at Birmingham (HP) and Aligning Science Across Parkinson’s [ASAP-020527] through the Michael J. Fox Foundation for Parkinson’s Research (MJFF) (HP, TRS, CFM, ZDW). For the purpose of open access, the authors have applied a CC BY public copyright license to all Author Accepted Manuscripts arising from this submission. Opinions, interpretations, conclusions, and recommendations are those of the authors and are not necessarily endorsed by the funding agencies.

## Author contribution

This is a collaborative team (genomics (HP), microbiology (TRS), statistics (CFM), bioinformatics (ZDW)). The lead/corresponding author (HP) conceptualized and developed the method. HP drafted the manuscript and all authors contributed to revision and approved the final version.

## Competing interest

HP has intellectual property pending on Personalized Microbiome-Based Therapies, Serial No.: 63/571,787; Filing Date: March 29, 2024. TRS is a co-inventor on intellectual property related to a small molecule microbiome-based therapy - US Patent 11,707,493 and 11,147, 792.

## Notes

### Author Declarations

The study used ONLY openly available human data that were originally published in Nat Comm (https://www.nature.com/articles/s41467-022-34667-x). The raw metagenomic sequences and accompanying metadata are located at the NCBI SRA under BioProject ID PRJNA834801 (https://www.ncbi.nlm.nih.gov/bioproject/834801), and post-QC and post taxonomic profiling data are on Zenodo (https://zenodo.org/record/7246185).

### Summary of Updates

The revision was made to (1) clarify that this is not a method for discovering disease associated features, in fact, having well-established disease-associated features are a requisite (2) to fulfill funding agency requirements on open access (3) include Zach Wallen as co-author.

